# Hypergraph Clustering for Analyzing Chronic Disease Patterns in Mild Cognitive Impairment Reversion and Progression

**DOI:** 10.1101/2025.04.14.25325825

**Authors:** Muskan Garg, Xingyi Liu, Maria Vassilaki, Ronald C Petersen, Jennifer St Sauver, Sunghwan Sohn

## Abstract

**Background:** Limited research has explored the preceding sequences of medical conditions leading to mild cognitive impairment (MCI), particularly patterns that may signal progression to dementia or reversion to normal cognition.

**Objectives:** Our study aims to analyze common *sequences* of chronic conditions preceding an MCI diagnosis and examine differences between retention in MCI or progression towards dementia and reversion to normal cognition, using hypergraph clustering, a network analysis approach.

**Methods:** We categorize participants into two groups (i) M2P: stay or progressed to dementia, or (ii) M2N: reversion to normal within 5 years after the first onset of MCI. Among 414 participants, 210 are males and the mean age is 80.8 years. We performed network analysis to obtain categories and sequences of chronic conditions. We applied hypergraph spectral clustering to characterize participants with similar sequences.

**Results: Results:** We identify and validate the generic key indicators (e.g., chronic kidney disease), highlight the sex-specific potential indicators (e.g., arthritis) for MCI reversal, and open new research directions for identifying potential disparities among men and women.

**Conclusion:** We categorized the chronic conditions preceding MCI diagnosis and discovered unique sequences suggesting MCI reversion among men and women to facilitate future research.

## 1. Introduction

The global prevalence of dementia is projected to triple by 2050. Around 6.2 million Americans aged 65 and above are diagnosed with Alzheimer’s dementia. Without significant medical advancements that can prevent, halt, or cure Alzheimer’s Disease (AD), this figure might double, reaching 13.8 million by 2060 (Nandi et al., 2024). This surge in numbers points to the importance of understanding and managing the condition more effectively, with a particular emphasis on its early stages and potential precursors of serious cognitive decline, such as mild cognitive impairment (MCI). MCI is characterized by noticeable changes in cognitive functions, including memory, decision-making, and language skills, which are more significant than what might be expected from normal aging but not severe enough to interfere with daily life (Lindbergh et al., 2016).

Understanding specific risk factors or chronic conditions that contribute to cognitive decline can aid healthcare providers in initiating early management strategies and monitor the state of cognition (Coon & Gómez-Morales, 2023; Gómez-Gardeñes & Moreno, 2006; Verghese et al., 2003; Wang et al., 2023; Zhang et al., 2022). MCI reversion, where previously diagnosed MCI returns to a state of normal cognition, suggests that the cognitive decline in the prodromal (pre-dementia) stage might not always be linear or inevitable (Bredesen et al., 2016; Rao et al., 2021), offering hope for interventions that could halt or even reverse this decline in its early stages. Recent longitudinal studies have identified potentially modifiable factors such as vision, arthritis, or mental ability (Koepsell & Monsell, 2012; Sachdev et al., 2013), quality of life (Overton et al., 2023; Trevisan et al., 2022), hemodialysis sessions (Li et al., 2023) and demographic factors (Overton et al., 2023; Yu et al., 2024) that influence the progression, stabilization, or reversal of MCI.

Existing research explores the relationship between chronic conditions and MCI trajectories, with diabetes identified as a risk factor for progression to dementia, while prediabetes and optimal glycemic control may facilitate MCI reversion. (Hu et al., 2020)(Makino et al., 2021). Cardiovascular conditions are associated with persistent MCI, though findings on their impact on reversion remain inconclusive (Welstead et al., 2021; Xue et al., 2019)(Sachdev et al., 2013).

However, existing research lacks a detailed analysis of the impact of pre-existing chronic conditions on the course of MCI, specifically in terms of its progression or potential reversal after diagnosis (McDowell et al., 2024; Yu et al., 2024). If specific patterns of chronic condition progression are associated with different post-MCI outcomes, they could serve as predictive markers, contributing towards personalized medicine. The existing longitudinal studies with demographic analysis (Hu et al., 2020; Makino et al., 2021; Sachdev et al., 2013; Welstead et al., 2021; Xue et al., 2019) revealed the need to identify associated patterns among chronic conditions and MCI reversal. Understanding MCI progression requires a shift in perspective, examining not just the factors contributing toward the progression of MCI into dementia but also the factors that might allow for a rollback to normal cognition (Overton et al., 2020).

To address this gap, we designed and developed an approach using hypergraph clustering to characterize the progression of chronic conditions. Our study addresses this gap by analyzing temporal patterns of chronic conditions and whether these patterns differ between those who revert from MCI to normal cognition (M2N) and those who stay at MCI or progress to dementia (M2P). We do not assert the contribution of specific chronic conditions to MCI progression or reversal but suggest the sequences linked to these outcomes. We collected chronic conditions of people from their health care visits spanning the five years before their MCI diagnosis and records of their cognition status in five years after MCI diagnosis as shown in Figure 1. We focussed on two aspects: (1) sequential occurrences of chronic conditions to identify notable patterns over time, and (2) network-based characterization of the cohort between M2N and M2P groups. We further examined sex disparities in the progression of MCI.

**Figure 1:**
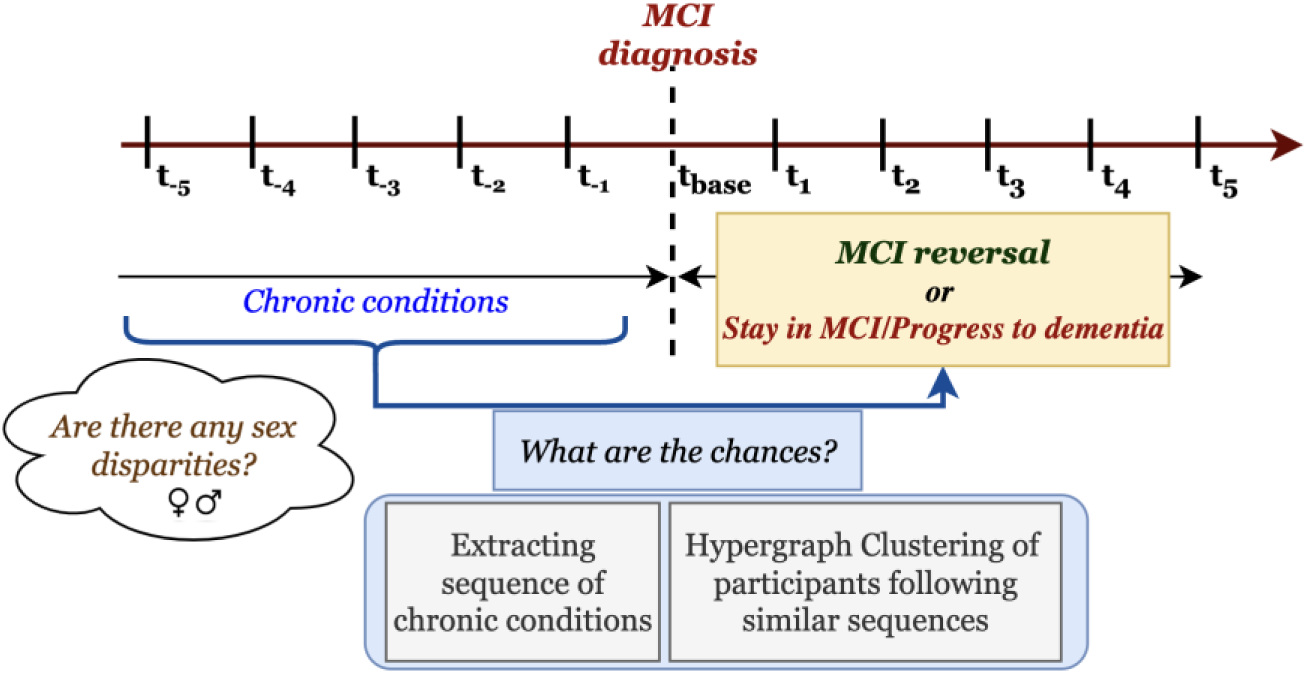
Overview of the study design

The research was approved by Mayo Clinic and Olmsted Medical Center Institutional Review Boards. Among 6,185, we selected participants from the Mayo Clinic Study of Aging (MCSA) cohort (Roberts et al., 2008) who were diagnosed with MCI, had at least one follow-up cognitive assessment within 5 years, and were diagnosed with at least one chronic medical condition within 5 years preceding MCI diagnosis. We obtained 414 participants. We extracted the ICD-9/10 codes of these participants from the Rochester Epidemiology Project (REP) infrastructure for 5 years before MCI diagnosis (St Sauver et al., 2012); and grouped them into clinical classification software (CCS) categories which were further classified into 15 chronic conditions recognized by the Department of Health, Housing, and Human Services as significant for aging (Goodman et al., 2013).

We tracked the sequence of newly added chronic conditions over five years preceding MCI and represented them in a network. After preprocessing and validating the network, we observed sequences, evolved from three types of chronic conditions (converging, diverging, and emerging). Aiming to discover the sequences of chronic conditions preceding MCI diagnosis, we removed the least frequent edges to obtain directed acyclic graphs and employed depth-first search algorithms to obtain the path in the network from a given source (chronic condition) to target (MCI). Finally, we found clusters of participants following similar sequences of chronic conditions and characterized them, highlighting the variations among different groups and age-specific stratification.

Our findings for chronic conditions sequence analysis and hypergraph spectral clustering suggest variation in progression among men and women; and examine differences between M2N and M2P.

## 2. Results

### Demographic characteristics of the study population

The participants were divided into two categories: (i) M2P (n=118): those who stayed on MCI or progressed to dementia within five years following their MCI diagnosis and (ii) M2N (n=296): those who reverted to a cognitively normal state within five years following their MCI diagnosis (see Table 1). Of 414 participants, 210 were male (50.8%) and 204 were female (49.2%) participants, with 253 persons (61.1%) aged over 80 years. Our primary interest lies in identifying the sequences of chronic conditions and characterizing participants based on the similarity of their trajectories including sex differences.

**Table 1.**
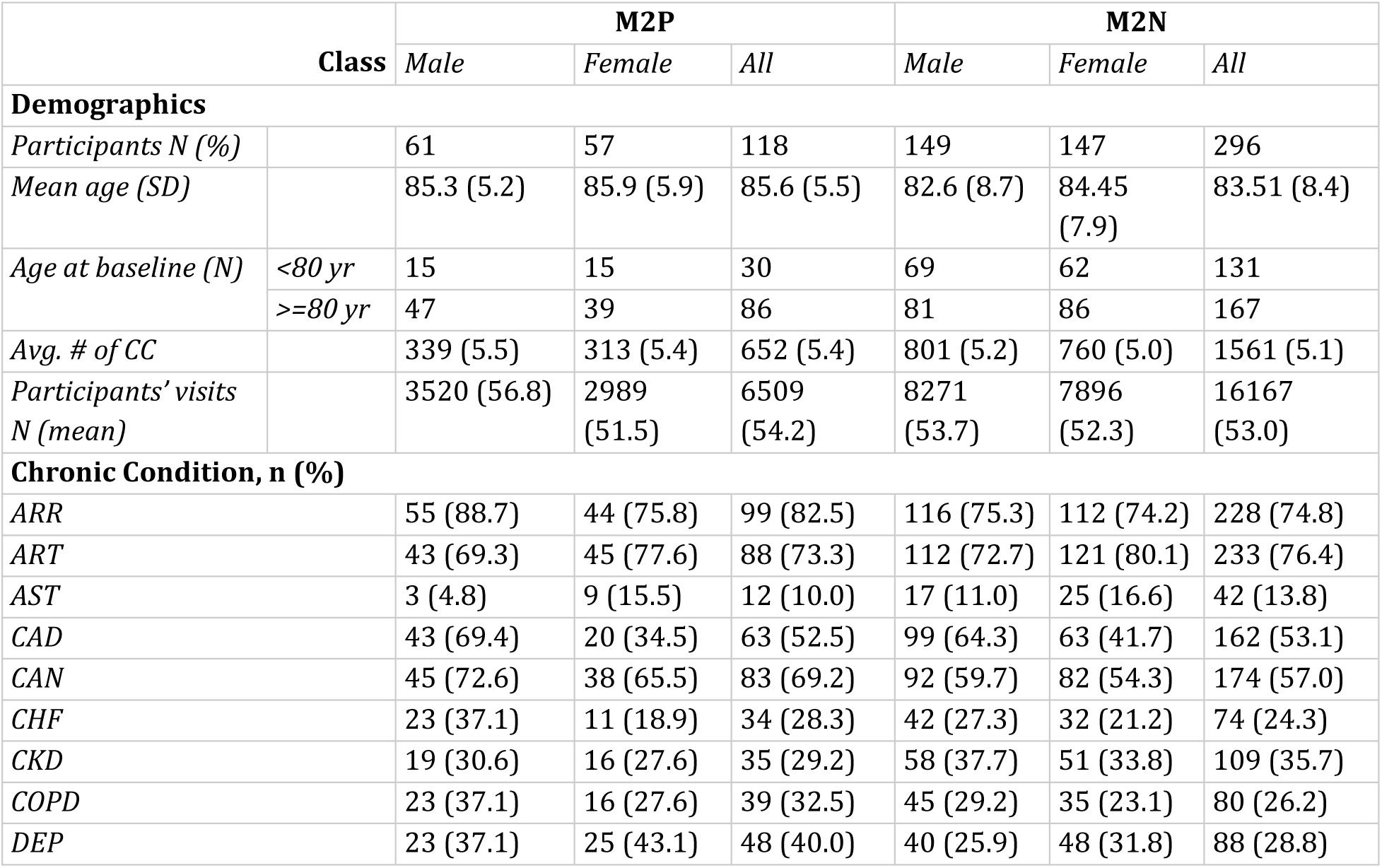
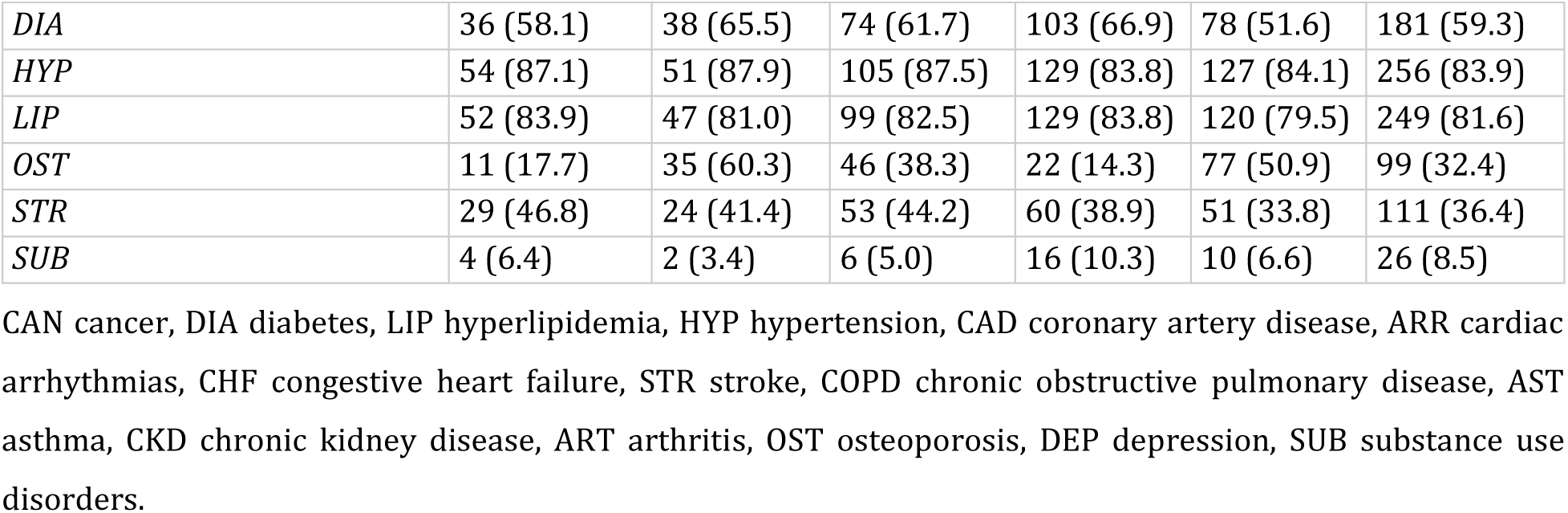
Characteristics of the study population: N number of participants, SD standard deviation, CC Chronic conditions.

| Class |  | M2P |  |  | M2N |  |  |
| --- | --- | --- | --- | --- | --- | --- | --- |
|  |  | Male | Female | All | Male | Female | All |
| **Demographics** |  |  |  |  |  |  |  |
| Participants N (%) |  | 61 | 57 | 118 | 149 | 147 | 296 |
| Mean age (SD) |  | 85.3 (5.2) | 85.9 (5.9) | 85.6 (5.5) | 82.6 (8.7) | 84.45 (7.9) | 83.51 (8.4) |
| Age at baseline (N) | <80 yr | 15 | 15 | 30 | 69 | 62 | 131 |
|  | >=80 yr | 47 | 39 | 86 | 81 | 86 | 167 |
| Avg. # of CC |  | 339 (5.5) | 313 (5.4) | 652 (5.4) | 801 (5.2) | 760 (5.0) | 1561 (5.1) |
| Participants' visits N (mean) |  | 3520 (56.8) | 2989 (51.5) | 6509 (54.2) | 8271 (53.7) | 7896 (52.3) | 16167 (53.0) |
| **Chronic Condition, n (%)** |  |  |  |  |  |  |  |
| ARR |  | 55 (88.7) | 44 (75.8) | 99 (82.5) | 116 (75.3) | 112 (74.2) | 228 (74.8) |
| ART |  | 43 (69.3) | 45 (77.6) | 88 (73.3) | 112 (72.7) | 121 (80.1) | 233 (76.4) |
| AST |  | 3 (4.8) | 9 (15.5) | 12 (10.0) | 17 (11.0) | 25 (16.6) | 42 (13.8) |
| CAD |  | 43 (69.4) | 20 (34.5) | 63 (52.5) | 99 (64.3) | 63 (41.7) | 162 (53.1) |
| CAN |  | 45 (72.6) | 38 (65.5) | 83 (69.2) | 92 (59.7) | 82 (54.3) | 174 (57.0) |
| CHF |  | 23 (37.1) | 11 (18.9) | 34 (28.3) | 42 (27.3) | 32 (21.2) | 74 (24.3) |
| CKD |  | 19 (30.6) | 16 (27.6) | 35 (29.2) | 58 (37.7) | 51 (33.8) | 109 (35.7) |
| COPD |  | 23 (37.1) | 16 (27.6) | 39 (32.5) | 45 (29.2) | 35 (23.1) | 80 (26.2) |
| DEP |  | 23 (37.1) | 25 (43.1) | 48 (40.0) | 40 (25.9) | 48 (31.8) | 88 (28.8) |

|  |  |  |  |  |  |  |
| --- | --- | --- | --- | --- | --- | --- |
| *DIA* | 36 (58.1) | 38 (65.5) | 74 (61.7) | 103 (66.9) | 78 (51.6) | 181 (59.3) |
| *HYP* | 54 (87.1) | 51 (87.9) | 105 (87.5) | 129 (83.8) | 127 (84.1) | 256 (83.9) |
| *LIP* | 52 (83.9) | 47 (81.0) | 99 (82.5) | 129 (83.8) | 120 (79.5) | 249 (81.6) |
| *OST* | 11 (17.7) | 35 (60.3) | 46 (38.3) | 22 (14.3) | 77 (50.9) | 99 (32.4) |
| *STR* | 29 (46.8) | 24 (41.4) | 53 (44.2) | 60 (38.9) | 51 (33.8) | 111 (36.4) |
| *SUB* | 4 (6.4) | 2 (3.4) | 6 (5.0) | 16 (10.3) | 10 (6.6) | 26 (8.5) |

CAN cancer, DIA diabetes, LIP hyperlipidemia, HYP hypertension, CAD coronary artery disease, ARR cardiac arrhythmias, CHF congestive heart failure, STR stroke, COPD chronic obstructive pulmonary disease, AST asthma, CKD chronic kidney disease, ART arthritis, OST osteoporosis, DEP depression, SUB substance use disorders.

### Classification of chronic conditions

In Figure 2, Graph G_top is the sample of a decomposed *network* of graph G evolved from progression of chronic conditions during five years preceding MCI diagnosis. In directed graph G_top, every node is a chronic condition, edge shows transition from one chronic condition to the other, and edge weight is the frequency of this transition. Due to high connectivity in the graph, we removed low-weighted edges to retain the most frequent transitions among chronic conditions. To resolve this, we apply k-bridge decomposition (Garg & Kumar, 2018), where k represents the minimum edge weight of every edge in the resulting graph. To determine the threshold value of k, we used the Pareto principle (i.e., roughly 80% of consequences come from 20% of causes) (Naoum et al., 2016), retaining the top 20% of the highest-weighted edges. We finally obtained a *decomposed network*.

**Figure 2:**
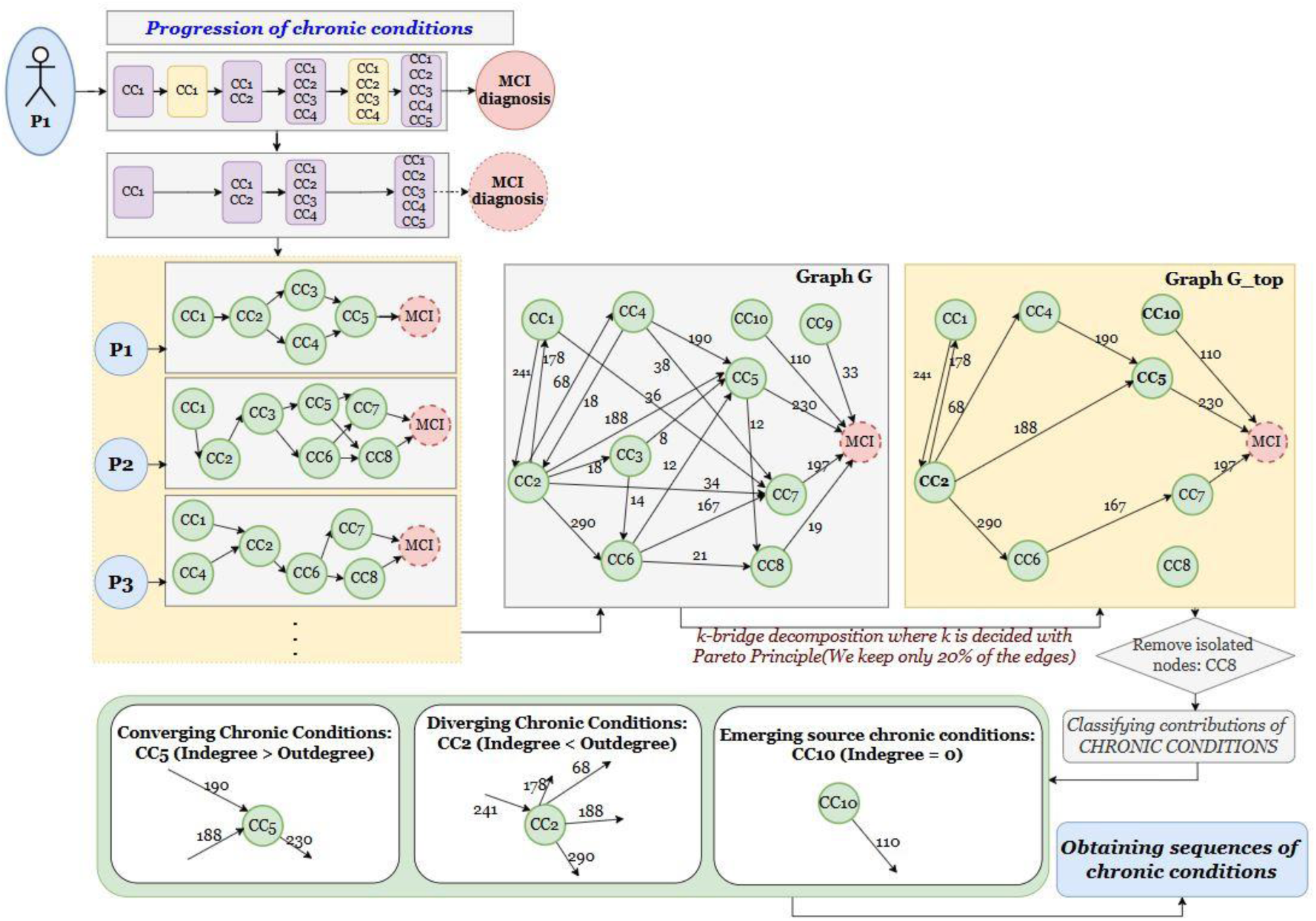
Extraction of chronic condition sequences: This figure illustrates the construction of a network and the classification of chronic conditions into converging, diverging, and emerging sources. CC denotes a chronic condition.

*Validating network of chronic conditions*: We validate the structural properties of the network for sex-stratified groups for M2N, M2P, or combined participants by comparing it with Erdős-Rényi (ER) (Gómez-Gardeñes & Moreno, 2006) and Watts-Strogatz (WS) models (Watts & Strogatz, 1998) and determine whether the network is random or structured. Table 2 contains the resulting values of characteristic path length (CPL) (Fronczak et al., 2004) and betweenness centrality (BC) (Barthelemy, 2004) of our graphs as compared to the other random networks. We observed almost ½ to 1/3^rd^ value of CPL in our graph as compared to ER model and WS model. In our graphs, shorter CPL indicates greater efficiency and higher interconnectivity in chronic condition transitions. The lower BC compared to random graphs—up to 4 to 6 times higher in the latter—suggests more alternative pathways, enhancing redundancy and reducing bottlenecks in disease progression. (see Table 2).

**Table 2:** Network metrics. Here, CPL is the characteristic path length, BC is the betweenness centrality, ER Erdos-Renyi model, and WS Watts-Strogatz model. The unit of values are the number of hops for CPL, and fraction of shortest paths for BC.

| Group |  | Our Graph |  | ER model |  | WS Model |  |
| --- | --- | --- | --- | --- | --- | --- | --- |
|  |  | CPL | BC | CPL | BC | CPL | BC |
| M2P | M | 1.06 | 0.04 | 1.17 | 0.02 | 1.33 | 0.04 |
|  | F | 0.42 | 0.01 | 1.55 | 0.06 | 1.46 | 0.04 |
|  | all | 0.77 | 0.02 | 1.51 | 0.05 | 1.53 | 0.05 |
| M2N | M | 0.80 | 0.02 | 1.43 | 0.04 | 1.45 | 0.04 |
|  | F | 0.79 | 0.02 | 1.52 | 0.04 | 1.53 | 0.05 |
|  | all | 0.69 | 0.01 | 1.41 | 0.04 | 1.50 | 0.05 |
| Both | M | 0.78 | 0.02 | 1.31 | 0.03 | 1.47 | 0.05 |
|  | F | 0.88 | 0.02 | 1.45 | 0.04 | 1.50 | 0.04 |
|  | all | 0.68 | 0.01 | 1.41 | 0.04 | 1.51 | 0.05 |

This is evident by the fact that multiple sequences of chronic conditions are important, suggesting complexities in trajectory. In the M2P group, the BC for graphs from men’s progression sequences resembles that of random graphs, indicating greater complexity for men. In contrast, women’s chronic conditions are more interconnected.

*Observing chronic conditions*: We removed the nodes with degree zero (isolated nodes) from the resulting graph. We define three types of nodes (chronic conditions) in the resulting graph: (i) converging chronic conditions, (ii) diverging chronic conditions, and (iii) emerging source chronic conditions.

1. **Converging chronic conditions (CCC)**: We consider the nodes (chronic conditions) of the graph whose indegree is greater than or equal to the out-degree as CCC. This pattern indicates that certain chronic conditions appear after many other chronic conditions, but comparatively fewer other conditions frequently follow these conditions. For instance, in Figure 2, the CC_5_ happens after CC_2_ and CC_4,_ suggesting a prospective connection of (CC_2_, CC_4_) and CC_5_, however, we do not claim the nature of association as correlation or causation. We compare the presence of such multiple associations with the predecessor against fewer associations with successors (such as MCI in this case)
2. **Diverging chronic conditions (DCC)**: Diverging Chronic Conditions are defined as nodes in a graph with an in-degree less than their out-degree. These conditions may emerge as medical concerns after fewer conditions but are predecessors to a greater number of chronic conditions. Similarly to CCC, we compare the number of connections before (1 association) and after (4 associations) CC_2_, suggesting the possibility of a higher number of sequences when the source condition is DCC.
3. **Emerging source Chronic conditions (ESCC)**: We defined ESCC as a set of all the chronic conditions with zero in degree in minimum one of the cases. For example, CC_10_ is directly followed by MCI, suggesting possibly minimal association of CC_10_ with other chronic conditions.

For each of three types of chronic conditions, we employ depth first search as a path finding algorithm.

### Sequences of chronic conditions

Analyzing sequences from the decomposed network, we evaluate the percentage of participants for each condition as they progress through different chronic conditions across female, male, and all participants (see Figure 3). Our observations are based on sequences followed by a minimum of 20 participants.

**Figure 3:**
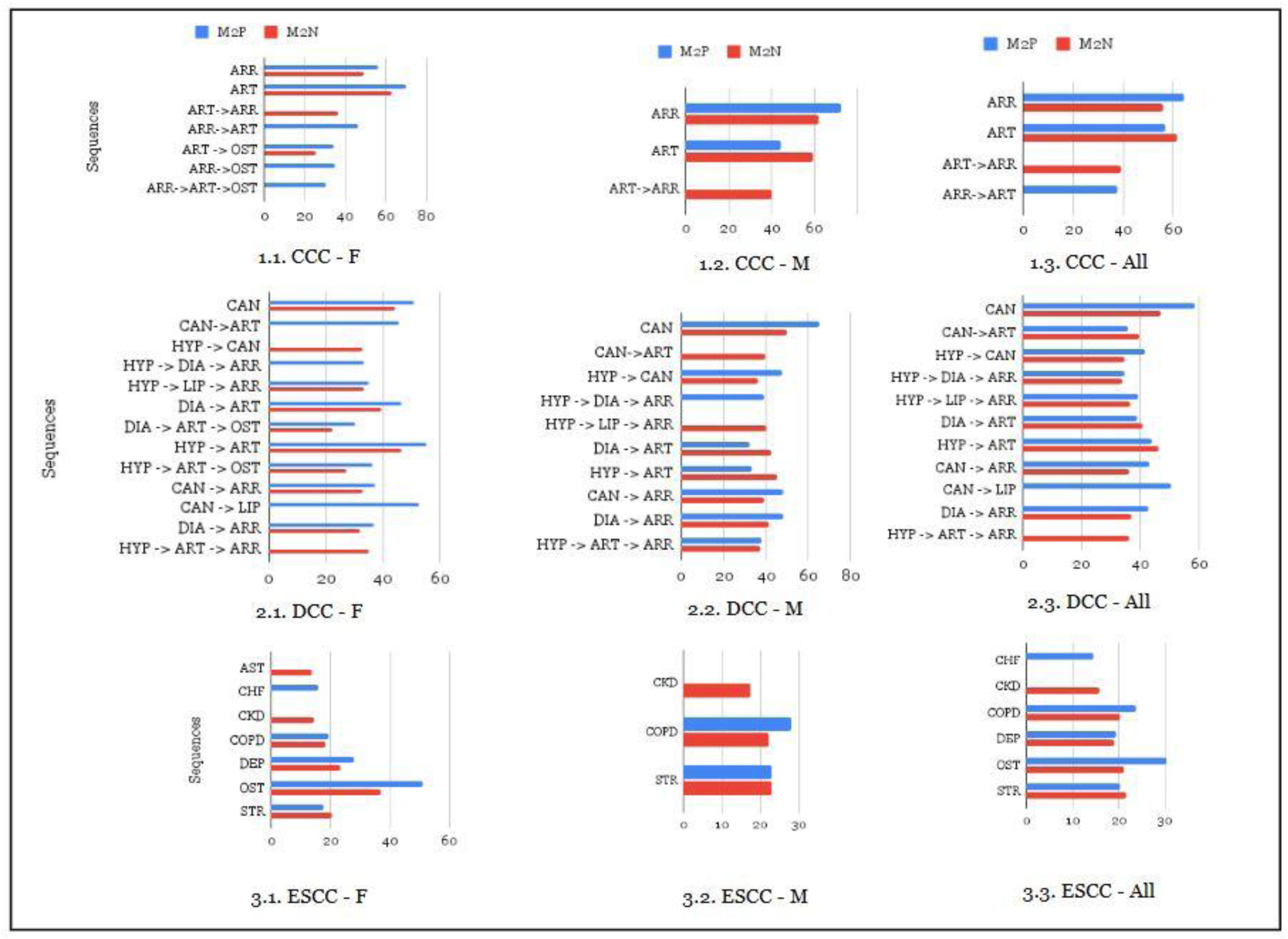
Experimental results of sequence analysis for *Converging Chronic Conditions (CCC), Diverging Chronic Conditions (DCC), and Emerging Source Chronic Conditions (ESCC) - Females (F), Males (M), and all participants (All).* Every sequence is followed by MCI. The x-axis represents the percentage of participants who follow a given sequence (y-axis) for the corresponding group - M2P (blue) or M2N (red). Sequences not among the top for a group are not shown in the graphs.

*Converging Chronic Conditions*: We found two chronic conditions as part of CCC, namely, ARR and ART. We obtained sequences evolving from these conditions to MCI diagnosis and observed differences among men and women. Among women, OST is observed as a potential M2P indicator, supported by research linking low bone density to higher dementia risk (Xiao et al., 2023). The sequence [‘ART → ARR → MCI’] is linked to MCI reversion, with a higher prevalence among women than men. Women with ARR may experience a more rapid cognitive decline than men, potentially impacting recovery (Volgman et al., 2019). Further research is needed to assess recovery rates, treatment prescriptions, progression timelines, and specific types of ART and ARR to guide targeted clinical interventions.

*Diverging chronic conditions (CAD, CAN, LIP, HYP, DIA)*: We observed the distribution of the percentage of observed sequences for DCC by the average number of participants in Appendix A. As expected, we observed a higher number of sequences using DCC. We ranked these sequences using the Analytical Hierarchical Process (AHP) optimization algorithm (Tavana et al., 2023) and obtained top sequences for sex-specific groups of participants. While the sequence [‘CAN → MCI’] suggests a higher likelihood of progression, [‘CAN → ART’] is associated with MCI reversion in males but progression in females. This highlights potential sex-specific differences in how cancer (CAN) and arthritis (ART) influence cognitive outcomes, warranting further investigation into biological, hormonal, and treatment-related factors. Although separate studies have examined the associations between (i) CAN and MCI (Ganguli, 2015) and (ii) ART and MCI (Weber et al., 2019), further research is recommended to explore sex-specific differences for effective interventions. The sequence [‘HYP → LIP → ARR’] links to MCI reversion in males but higher progression in females and the overall group, suggesting sex-specific mechanisms. Understanding these differences can aid in developing targeted interventions.

Notably, most sequences ending with [’… -> ART -> MCI’] show a higher likelihood of progression among female participants, whereas they exhibit a greater probability of reversion among male participants. This disparity may be driven by factors such as inflammation, hormonal differences, and comorbid conditions. Certain sequences, such as [’DIA → ART → MCI’] and [’HYP → ART → MCI’], exhibit significant sex-based differences, showing progression in females but MCI reversion in males. Managing hypertension and diabetes is crucial for cognitive health (Newby & Garfield, 2022); further analysis of their severity and timeline is needed to link them to MCI reversion.

*Emerging source chronic conditions (CHF, COPD, STR, DEP, AST, CKD, STR):* Most of the ESCC have out-degree one, suggesting the only connection, which is observed as MCI. We calculated the percentage of participants transitioning from emerging source chronic conditions to MCI. We observed that CKD participants recovered from MCI to normal cognition. Most concerning chronic conditions among women are DEP, and OST, indicating higher chances of M2P. Men exhibit a significantly lower frequency of hospital visits for conditions such as asthma (AST), osteoporosis (OST), and depression (DEP) compared to women We further analyzed the number of sequences covered by CCC, DCC and ESCC. DCC, due to its divergence, exhibits a significantly higher number of sequences (>150) compared to CCC and ESCC, which each display fewer than 20 sequences. This is probably due to a greater number of successor chronic conditions as compared to the predecessor chronic conditions. Thus, medical conditions such as CAD, CAN, LIP, DIA, and HYP combine in multiple combinations among themselves to form sequences. Hence, we noticed the complexities added by DCC while constructing a network from the sequence of chronic conditions followed by participants during hospital visits.

### Characterizing participants

We obtain clusters for each group through hypergraph spectral clustering and represent them for different types of chronic conditions (see Figure 4). In Figure 4, the dotted lines represent the ideal distribution, while the solid bubbles represent the actual distribution of participants in each cluster. Additionally, we observed the cluster distribution stratified by age for men, women, and all participants. We noticed most variations in M2P to M2N distribution and made observations for (i) MCI reversion, and (i) stay in MCI or progression to dementia.

**Figure 4:**
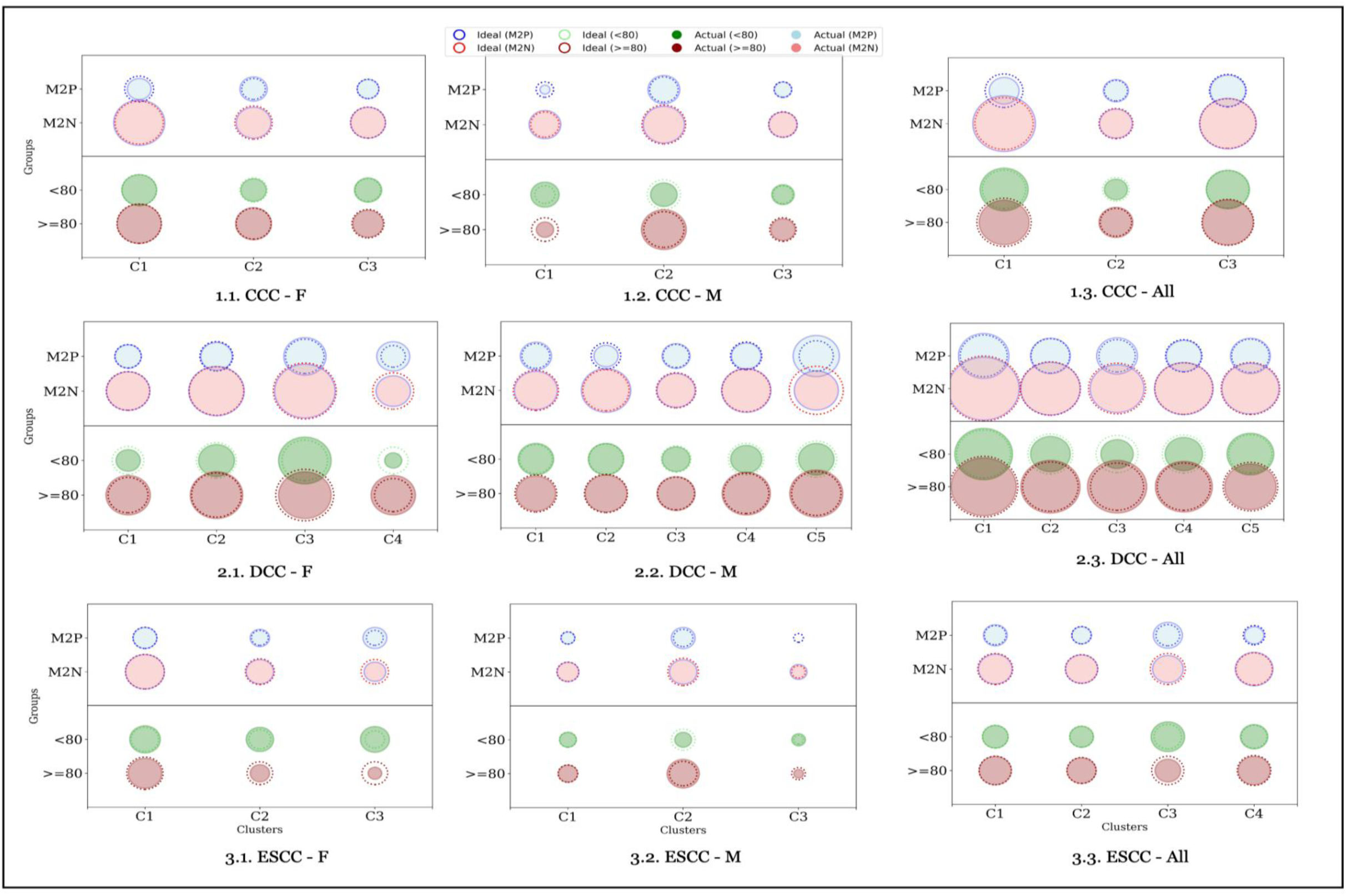
Characterizing participants with similar chronic condition sequences for *Converging Chronic Conditions (CCC), Diverging Chronic Conditions (DCC), and Emerging Source Chronic Conditions (ESCC) - Females (F), Males (M), and all participants (All).* Size of the bubble corresponds to the number of participants in each cluster from minimum (ESCC-M (C3): 3) to maximum (DCC-All (C1): 86). The clusters for each observation are different. For instance, C1 for females may not be the same as C1 in males.

*CCC Clusters*: In Figure 4, we considered the clusters with a minimum of 20 participants and observed the MCI reversion in cluster C1 of CCC- [F, All], and C2 of DCC-M. The sequences in cluster C1 for all participants are [’ART -> MCI’,’ART -> ARR -> MCI’]; and an additional sequence in C1 for females is [’ART -> OST -> MCI’]. Among C1 for all participants, the expected number of participants for M2P and M2N were 15 and 39, respectively. However, 40% of this number for M2P – 6 more participants – were found in M2N. This indicated that there are higher chances of reversal if participants follow the sequences ‘ART’ -> {*ϕ*, OST, ARR}-> ‘MCI’. An additional sequence in C1 for CCC-F is specifically associated with females and suggests a higher prevalence of OST among women, supporting our previous observations in Figure 3.

As we observe the clusters C2 for CCC-F^1^, CCC-M^2^, and CCC-All^3^, we find the sequences starting with ‘ARR’, suggesting an increase in the number of participants progressing towards cognitive decline. Thus, our observations indicate that the participants, following Arthritis (ART) -> (chronic conditions) -> MCI have a higher chance (the error rate of +16.1%) of reversal whereas the (ARR) -> (chronic conditions) -> MCI have a higher chance of progression (the error rate of +34.6%). Further investigations are required for association, correlation, and causal analysis.

These findings align with the patterns observed in Figure 3 (1.1-1.3), where’ARR’ ->’ART’ indicates progression, while’ART’ ->’ARR’ suggests MCI reversal. However, cluster 2, which contains sequences starting with’ARR’, includes a higher proportion of participants over 80 years old.

DCC Clusters: The cluster C2 for DCC-M contains multiple sequences^4^ evolving from hyperlipidemia and hypertension to MCI. Being divergent, HYP and LIP precede many chronic conditions. Although there is not much variation for other clusters of DCC, we noticed an 11.40% increase for M2N among 31 participants of cluster C2 in DCC-M. However, due to the presence of many sequences, it is difficult to determine which sequences are the cause of this reversal. In contrast to reversal patterns, participants in clusters for the sequences evolving with CAN: C4^5^ for DCC-F and C3^6^ for DCC-All, and the sequences evolving with LIP: C5^7^ for DCC- are more likely to experience progression with an error rate of +72.3%, +80.2%, and +29.4%, respectively. These observations highlight the impact of multimorbidity on cognitive decline, and most older participants are following these sequences.

ESCC Clusters: While cluster 3 of ESCC- [F, M, and All] exhibits considerable variation, the number of participants in these clusters is fewer than 20. Therefore, no definitive claims or observations can be drawn from this data.

## Discussion

We analyzed how the patterns of chronic conditions differ in M2P vs. M2N groups (stayed or progressed to dementia vs. reverted to normal) leveraging sequence analysis and hypergraph clustering. To the sequences from time-varying data, it is important to accommodate the progression of chronic conditions both sequentially and parallelly. As we observed the chronic conditions, we noticed the addition of more than one chronic condition in a single visit that are treated as parallel addition. Existing sequential pattern mining algorithms are limited to sequential patterns. By accommodating both sequential and parallel chronic conditions, we considered the complexities in the progression of chronic conditions leading to MCI diagnosis. Our network-based sequence mining method categorized chronic conditions into CCC, DCC, and ESCC, leveraging a classified approach.

After sequence analysis, we employed *hypergraph spectral clustering* for participant characterization. We define the problem as each sequence can be followed by multiple participants (M) and each participant can follow multiple sequences (N). This M to N mapping of sequences and participants is a bipartite problem which is represented as a hypergraph. The hypergraph spectral clustering allows clustering on the bipartite nature of the problem, unlike clustering of pairwise association in a conventional network. We clustered the sequences and accommodated the corresponding participants for calculating the chances of MCI reversal.

In our study cohort of 414 participants, 296 participants (71.5%) reverted from MCI to normal cognition in 5 years post-MCI diagnosis, suggesting that most subjects have reversed back to normal at least once after MCI diagnosis. As we considered the prevalence of chronic conditions in Table 1, we observed a higher frequency in the occurrence of ARR, LIP, and HYP among both men and women. However, a few chronic conditions are more frequent among men (CHF, CAD) and a few others in women (OST, AST). We observed no statistically significant difference among prevalent frequencies of M2P and M2N after applying the t-test. Among 15 chronic conditions considered for this study, we noticed the least frequency for AST and SUB, resulting in their least contribution towards analyses of progression.

We assess the structural properties of the resulting networks for three types of chronic conditions (converging chronic conditions (CCC), diverging chronic conditions (DCC), and emerging source chronic conditions (ESCC)) across female, male, and all participant groups.

The observed consistency in structural properties, along with their deviation from a randomly generated graph, indicates that the constructed networks encapsulate meaningful and nontrivial information about chronic condition progression. Our observations are based on sequence clusters with at least 20 participants.

We analyzed the number of sequences covered by CCC, DCC, and ESCC, with DCC showing the highest sequence diversity due to its greater number of successor conditions as shown in Figure 5(i). As expected, DCC covered the highest number of participants and sequences, followed by CCC, which included more participants but fewer sequences than ESCC as shown in Figure 5(ii). Furthermore, to understand the sequences and their association with participants, followed by characterizing participants, we employed hypergraph spectral clustering.

**Figure 5:**
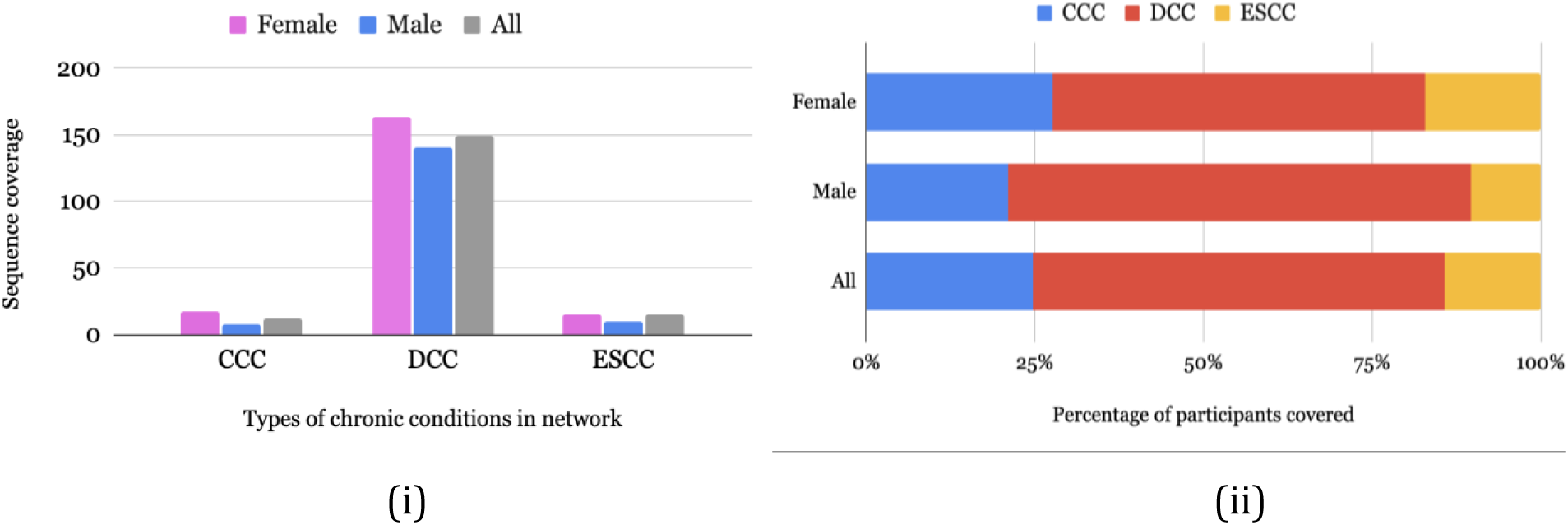
Coverage by CCC, DCC and ESCC: (i) sequences, and (ii) participants.

Our observations indicate that the sequences following ART -> (converging chronic conditions) -> MCI have higher chances of MCI reversal. The older participants (80+ years) in the former sequence show a higher tendency for progression, likely due to age-related vascular and inflammatory factors. Age-stratified studies and biomarker research are essential for understanding how arthritis impacts post-MCI cognitive status. Additionally, analyzing disease severity and timing can help guide effective clinical interventions. Moreover, sequences originating from arthritis (ART) are more commonly observed in women. These observations are more likely because of the later emergence of certain ART conditions in men compared to women. Given the diversity of arthritis types—where osteoarthritis is primarily associated with aging, while rheumatoid arthritis is not age-related (Serhal et al., 2020)—further investigation is necessary to assess the type and severity of arthritis for the development of targeted medical interventions. Previous studies emphasize the need for research at both the mechanistic and population levels to explore associations between rheumatoid arthritis and MCI (Meade et al., 2018; Sharma & Chen, 2023). Additionally, while an association between osteoarthritis and dementia risk has been observed, causality remains unestablished. This underscores the need for prospective cohort studies to determine whether knee, hip, or hand osteoarthritis independently contribute to cognitive decline (Weber et al., 2019).

We observed that fewer chronic conditions lead to DCC, but many follow it, contributing to further health complications. Most sequences in DCC clusters are linked to a higher-than-expected percentage of older individuals (>80 years), highlighting chronic conditions that become more prevalent in later life. Multiple diverging conditions, such as HYP, LIP, and CAN, lead to various chronic conditions, making it difficult to identify clear patterns. Additionally, most clusters in DCC contain multiple sequences, limiting meaningful or quantifiable observations. Cancer diagnosed just before MCI has a higher chance of MCI reversal compared to cases where cancer is followed by chronic conditions before MCI. This may be because the latter sequence is more common in older participants than younger ones. A careful analysis is required for the progression of chronic conditions post-cancer diagnosis especially those having HYP or LIP. The sequence of cancer followed by arthritis leading to MCI suggests reversion in men but progression in women, requiring further investigation for men and women.

As per cross-sectional analysis, COPD, CAN, and DEP indicate progression of cognitive decline post MCI diagnosis whereas ART and CKD show chances of MCI reversion. Research indicates that CKD treatments such as kidney transplantation (Viggiano et al., 2020) may support MCI reversal, suggesting a neuroprotective effect. Kidney transplantation enhances cerebral perfusion, reduced uremic toxin burden, and correction of metabolic imbalances (Pépin et al., 2023; Yan et al., 2024). Our ESCC cluster findings align with this, reinforcing the potential reversibility of CKD-related cognitive impairment. We found that DEP and OST are more likely to accelerate cognitive decline in women, likely due to neuropsychiatric and inflammatory pathways Our observations align with previous studies indicating that women are at a higher risk of late-life depression, which plays a significant role in the progression of dementia (Amouzougan et al., 2017; Ly et al., 2021; Sundermann et al., 2017). Chronic depression has been associated with increased β-amyloid deposition and impaired executive function (Li et al., 2022), highlighting the need to investigate relevant biomarkers to enhance medical interventions. Additionally, research indicates that osteoporosis (OST) leads to reduced bone density, and individuals with low bone density are at a higher risk of developing dementia (Curtis et al., 2024; Xiao et al., 2023). However, we do not establish causality for MCI reversion. Further analysis is needed to understand the lower diagnosis rates of certain ESCC conditions (AST, DEP, and OST) among men, which may be influenced by cultural attitudes, lower symptom awareness, and underutilization of preventive care. Our findings show that ESCC sequences leading to MCI diagnosis differ in frequency between M2N and M2P groups and in occurrence between men and women. A deeper investigation is required to determine whether men are less concerned about these conditions or less likely to seek treatment.

For progression-related sequences, targeted medical interventions should focus on early management of cancer, hyperlipidemia, and hypertension to mitigate cognitive decline, with special attention to osteoporosis and depression in women. One of the key observations across notable sequences for MCI reversal is the increase in the number of younger participants (<80 years) than expected. Age significantly influences cognitive decline, with older individuals more prone to MCI progression due to ARR, arthritis, and vascular issues. Younger individuals or those with higher cognitive reserve may show better resilience, potentially stabilizing or improving cognition. Age-stratified studies are needed to explore how ARR and ART impact MCI progression. Higher brain reserve can delay cognitive decline, while lower reserve or genetic factors may accelerate it. Biomarker studies are crucial for further understanding. Examining variations in arrhythmia and arthritis severity and timing may aid clinical interventions.

*Limitations*: We carry forward chronic conditions while omitting the non-chronic conditions to mitigate potential distortions or anomalies caused by isolated incidents that may or may not occur in conjunction with cognitive decline. We did not consider the treatments administered or the severity of the conditions at this stage of our analysis. Although our findings have potential implications for improving patient care, the primary contribution of our study is providing a methodological framework for analyzing longitudinal health conditions. The generalizability of these findings needs validation in other clinical settings.

## 4. Conclusion

Our hypergraph clustering approach uncovers unique chronic condition sequences from longitudinal healthcare visits leading to MCI diagnosis. We highlight three categories of chronic conditions preceding MCI diagnosis and reveal their structural properties to facilitate future research. We identify distinct sequences linked to the stay or progression of MCI and MCI reversion, encouraging a deeper exploration of sex-and age-specific disease progression.

## 5. Method

### Study setting and design

The MCSA is an ongoing population-based study focused on understanding cognitive aging, conducting detailed cognitive assessments on participants every 15 months. Individuals from Olmsted County, Minnesota, were chosen through a randomized process that considered age and sex, ensuring a diverse representation. Each participant underwent a thorough evaluation by three independent assessors: a study coordinator, a physician, and a psychometrist. A diagnosis for each participant, whether MCI, dementia, or no cognitive impairment, was made through consensus by a committee consisting of three evaluators (including a neuropsychologist) (Roberts et al., 2008).

In our study, we used the REP data to extract the ICD codes of the chronic conditions of the MCSA participants who were diagnosed with MCI. The REP is a medical research infrastructure established in the early 1960s and encompasses the medical records of residents in Olmsted County and other 27 counties across Minnesota, Wisconsin, and Iowa.

The key feature of the REP is its ability to link together the medical records from different healthcare providers in the community, including hospitals, clinics, and private practices (St Sauver et al., 2012). This linkage allows researchers to follow individuals over time and across different healthcare settings, providing a longitudinal view of their medical history.

### Network construction

We first obtained the MCSA participants’ chronic conditions over the period of five years before MCI diagnosis. Since chronic conditions are added in subsequent visits, we keep track of the newly added chronic conditions. We constructed a network, depicting the progression of chronic conditions as a state diagram. We merged these state diagrams as a single network, the event-based (occurrence of chronic conditions) representation among participants. Thus, graph G is constructed by merging the state transition diagrams (evolved from the progression of chronic conditions) for each participant.

The graph evolved from top sequences of chronic conditions can reveal common trajectories of progression, facilitating future research for discovering comorbidities, and potential causative relationships between conditions. Unlike random graphs, which might show no meaningful patterns or relationships, the resulting graph can highlight statistically significant patterns and trends that have real-world implications on progression of cognitive decline. We evaluated the nature of the observed graph against random models to demonstrate the non-random nature of the chronic condition sequences using average shortest path length and betweenness centrality. An Erdos-Renyi graph connects each pair of nodes (chronic conditions) with a fixed probability p, serving as a benchmark for understanding random connectivity. A Watts-Strogatz graph is constructed to capture the small-world properties observed in many real-world networks. It starts with a regular lattice and then re-wires edges randomly with a probability p, creating a mix of local clustering and short global paths. In this work, we investigate the network topology in observed and random graphs to determine how chronic conditions are spread throughout the network, uncovering the significance of underlying patterns in the observed network.

The nature of a graph evolved from a sequence of nodes, is often tested with characteristic path length and betweenness centrality to gain insights into its structure and dynamics. The characteristic path length (CPL) of a network is the average shortest path length between all pairs of chronic conditions. It provides a measure of the typical distance between chronic conditions. Betweenness centrality (BC) measures the extent to which a chronic condition lies on the shortest paths between other chronic conditions. It quantifies the significance of chronic conditions as intermediaries in the network.

### Path finding

We applied depth-first search for each of three types of nodes (chronic conditions) in the resulting graph: (i) Converging chronic conditions (CCC), (ii) Diverging chronic conditions (DCC), and (iii) Emerging source chronic conditions (ESCC). The resulting sequence list represents the possible combinations of chronic conditions by preserving the temporal order. The primary goal of constructing these sequences is to analyze the progression and emergence of chronic conditions over specified intervals for participants. We considered separate cases for groups: (i) group 1 – M2P, (ii) group 2 – M2N, and (iii) group 1 and 2 and observed filtered networks for (a) male, (b) female, and (c) all participants for each of these groups. After applying these permutations, we obtain 3 ∗ 3 = 9 different cases which are referred to as cases in the remaining part of the manuscript. We obtain sequences by finding a path from an element in each type of chronic condition as the source node and ‘MCI’ as the target node. First, we identified cycles in the graph and removed the least weighted edge to obtain the directed acyclic graph (DAG) (Digitale et al., 2022). We employed the depth-first search (DFS) (Tarjan, 1972) on DAG to obtain the path obtained as sequences of any length greater than or equal to two. We examined the number of sequences and coverage of the number of participants in CCC, DCC, and ESCC.

### Hypergraph Clustering

A hypergraph naturally captures many-to-many relationships where a characteristic is shared among multiple entities (nodes). This is particularly useful in health-related studies where disease progression can be similar across a group of individuals (Cai et al., 2022; Somu et al., 2016). By using hyperedges to represent these relationships, we can easily analyze clusters of people with similar health patterns, potentially uncovering shared risk factors or outcomes that might not be as apparent when only looking at pairwise interactions. Thus, an edge in the hypergraph is defined as a *hyperedge* that can connect any number of vertices.

If participants *A,B and C* all follow the same sequence of chronic conditions (e.g., diabetes followed by hypertension and then arthritis), these participants would be collectively connected by a single hyperedge in the hypergraph. This connection reflects their shared medical trajectory, which is more complex than pairwise relationships and might indicate a common underlying health profile. We proceeded to extract clusters using hypergraph spectral clustering (Michoel & Nachtergaele, 2012) to identify subgraphs with groups of participants that follow similar chronic condition sequences as shown in Figure 6.

**Figure 6:**
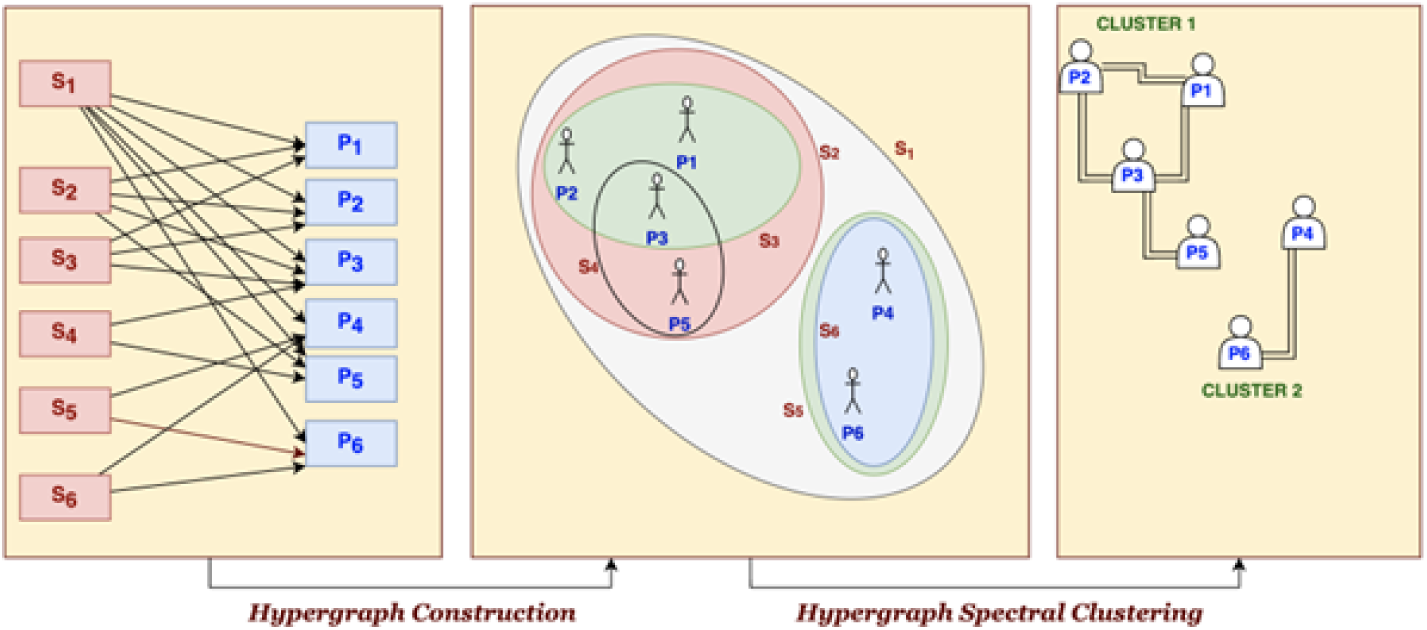
Overview of Hypergraph analysis. S_i_ denotes i^th^ sequence and P_j_ denotes j^th^ participant. Hypergraph Spectral Clustering is a method used in data analysis and machine learning to partition hypergraphs into clusters based on the spectral properties of their associated matrices. It extends the concept of spectral clustering from traditional graphs to hypergraphs. We employ Algorithm 1 for hypergraph spectral clustering and analysis.

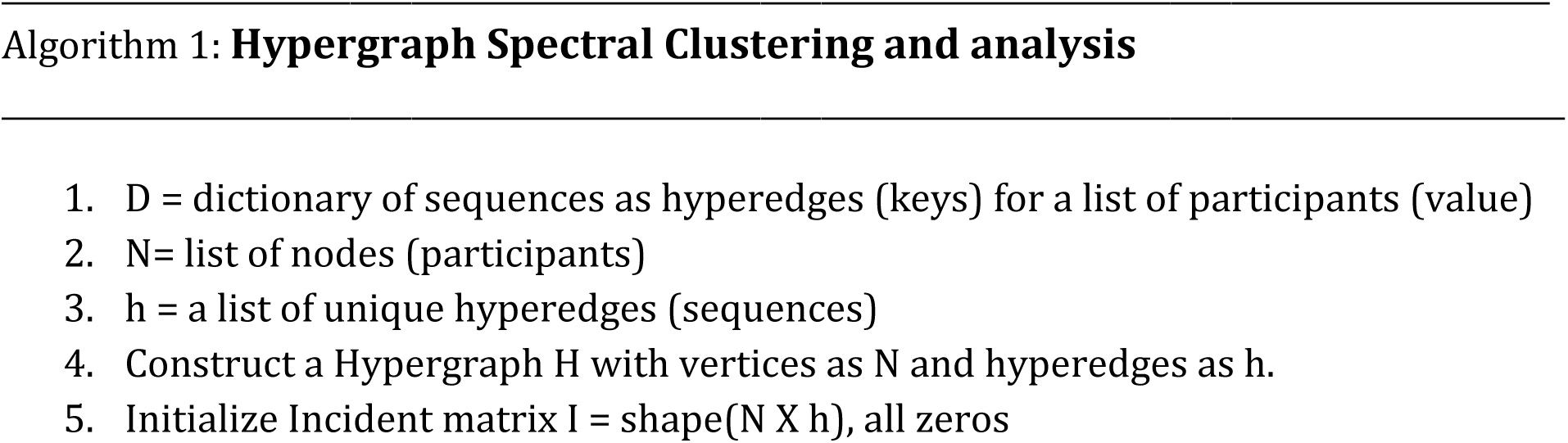
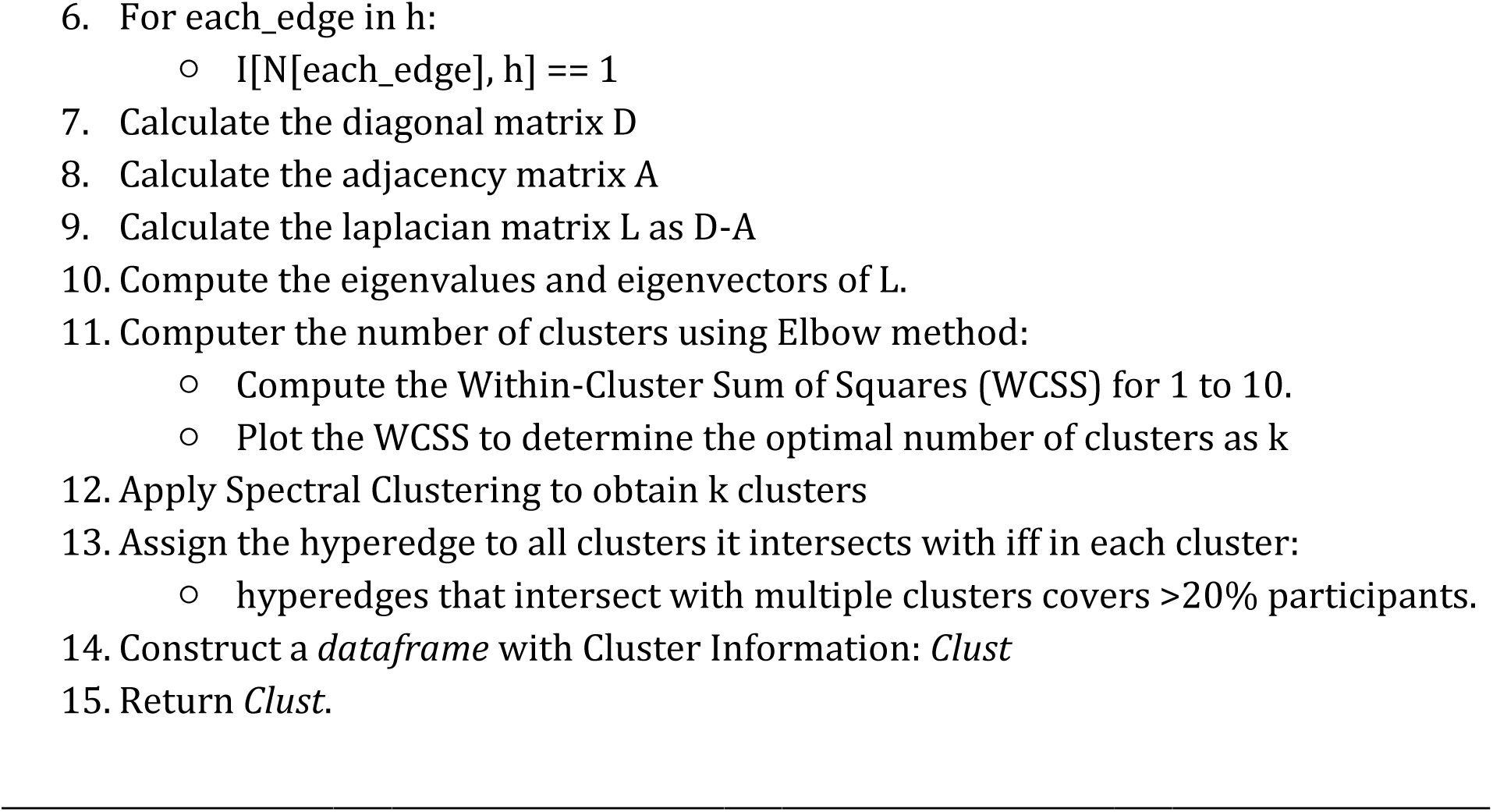

Multi-core processors (e.g., Intel i7/i9, AMD Ryzen) are essential for efficient parallel computations. A minimum of 8 GB of RAM is required for moderate-sized networks, with 16 GB or more recommended for larger datasets or more complex analyses. For our analysis, we used Python along with several libraries: NetworkX for network analysis and sequence extraction, scikit-learn and NumPy for numerical operations and linear algebra in spectral clustering, HyperNetX for working with hypergraphs, and Matplotlib for visualizing results.

After finding the clusters with corresponding participants, we quantify deviation among participants, characterized by sex-specific and age-specific ratios. Consider the number of participants in the original data as *N_o_*.

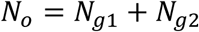

Where *N_g1_* is the number of participants belonging to M2P group and *N_g2_* is the number of participants belonging to M2N group. The *base ratio of participant count* is denoted as *N_g1_*: *N_g2_*. The base ratio suggests that the participants should be in the same ratio of groups in the resulting cluster. Similarly, we considered the ratio of the number of participants for age-specific characterization.

Consider a set of clusters obtained as C and each cluster *C_i_* has coverage of *N_i_*. Coverage refers to the number of participants who adhere to the patterns or sequences found in the resulting cluster *C_i_* from hypergraph spectral clustering. We divide *N_i_* according to the base ratio *N_g1_*: *N_g2_* which gives the expected number of participants in each group as

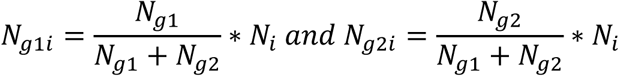

Here, *N_g1i_* and *N_g2i_* are the expected numbers of participants who belong to groups 1 and 2, respectively, within the cluster *C_i_* and we referred them as expected values (EV). The actual observed counts of participants in group 1 and group 2 in *C_i_* are recorded as observed values (OV). These observed values are compared to the expected values to assess the distribution within the cluster through percentage error.

Percentage Error quantifies how much the observed value deviates from the expected value, expressed as a percentage of the expected value. It shows the extent of increase or decrease in the number of participants relative to what was expected. It is defined as:

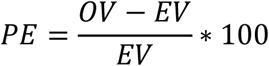

Where PE is the percentage error, OV is the observed value, EV is the expected value.

## Data Availability

The MCSA study makes de-identified data available to qualified researchers upon reasonable request.

## Footnotes

1 [’ARR -> OST -> MCI’,’ARR -> ART -> MCI’,’ARR -> MCI’]

2 [’ARR -> MCI’,’ARR -> ART -> MCI’]

3 [’ARR -> MCI’]

4 [’LIP -> MCI’,’HYP -> ART -> ARR -> MCI’,’HYP -> ART -> MCI’,’HYP -> ARR -> MCI’,’HYP -> CAD -> MCI’,’HYP -> LIP -> MCI’,’HYP -> CAN -> MCI’,’HYP -> MCI’,’HYP -> DIA -> MCI’,’HYP -> CKD -> MCI’,’LIP -> ARR -> MCI’,’LIP -> CAN -> MCI’,’LIP -> DIA -> MCI’,’LIP -> ART -> MCI’,’LIP -> CAD -> MCI’]

5 [’CAN -> HYP -> OST -> MCI’,’CAN -> HYP -> DIA -> MCI’,’CAN -> HYP -> ART -> MCI’,’CAN -> HYP -> MCI’,’CAN -> ARR -> ART -> MCI’,’CAN -> ARR -> MCI’,’CAN -> LIP -> ART -> MCI’,’CAN -> LIP -> MCI’,’CAN -> OST -> MCI’,’CAN -> ART -> MCI’,’CAN -> MCI’,’CAN -> HYP -> LIP -> MCI’,’CAN -> HYP -> ARR -> MCI’]

6 [’CAN -> ARR -> MCI’,’CAN -> LIP -> ART -> MCI’,’CAN -> LIP -> MCI’,’CAN -> DIA -> ART -> MCI’,’CAN -> DIA -> MCI’,’CAN -> ART -> MCI’,’CAN -> MCI’,’CAN -> LIP -> ARR -> MCI’]

7 [’CAN -> ARR -> MCI’,’CAN -> DIA -> MCI’,’CAN -> MCI’,’DIA -> MCI’,’CAN -> ART -> MCI’]

## Acknowledgements

This study was supported by NIH (National Institutes of Health) R01 AG068007. The Mayo Clinic Study of Aging was supported by NIH Grants (U01 AG006786, P30 AG062677, R37 AG011378, R01 AG041851, R01 NS097495), the Alexander Family Alzheimer’s Disease Research Professorship of the Mayo Clinic, the Mayo Foundation for Medical Education and Research, the Liston Award, the GHR Foundation, the Schuler Foundation, and used the resources of the Rochester Epidemiology Project (REP) medical records linkage system, which is supported by the National Institute on Aging (NIA: AG 058738), by the Mayo Clinic Research Committee, and by fees paid annually by REP users.

## Authors’ contributions

M.G. conceptualized and designed the study, developed the model, collected, analyzed, and interpreted the data, drafted the initial manuscript, and revised the manuscript. M.V. and J.S. analyzed and interpreted the data and revised the manuscript. X.L. and R.P. interpreted the data and revised the manuscript. S.S conceptualized and designed the study, supervised data collection, model development, and analysis, interpreted the data, and reviewed and finalized the manuscript. All authors approved the final manuscript and agreed to be accountable for all aspects of the work.

## Data availability statement

The MCSA study makes de-identified data available to qualified researchers upon reasonable request.

## Ethics approval and consent to participate

This study was approved by the Mayo Clinic Institutional Review Board and the Olmsted Medical Center Institutional Review Boards. Patients included in this study provided informed consent at the time they enrolled in the Mayo Clinic Study of Ageing (MCSA). We have not developed questionnaire and interview for this study.

## APPENDIX A

We observed the maximum sequence follow-up by 20 to 40 percent of the participants. However, there is a low percentage of sequences followed by more than 40% of the participants as marked by a dotted green line in Figure 7 (a-c). We further make observations for M2N and M2P. We observed high overlapping sequence distribution for M2N and M2P for males (see Figure 7 (b)), unlike sequence distribution for women (see Figure 7 (c)).

**Figure 7:**
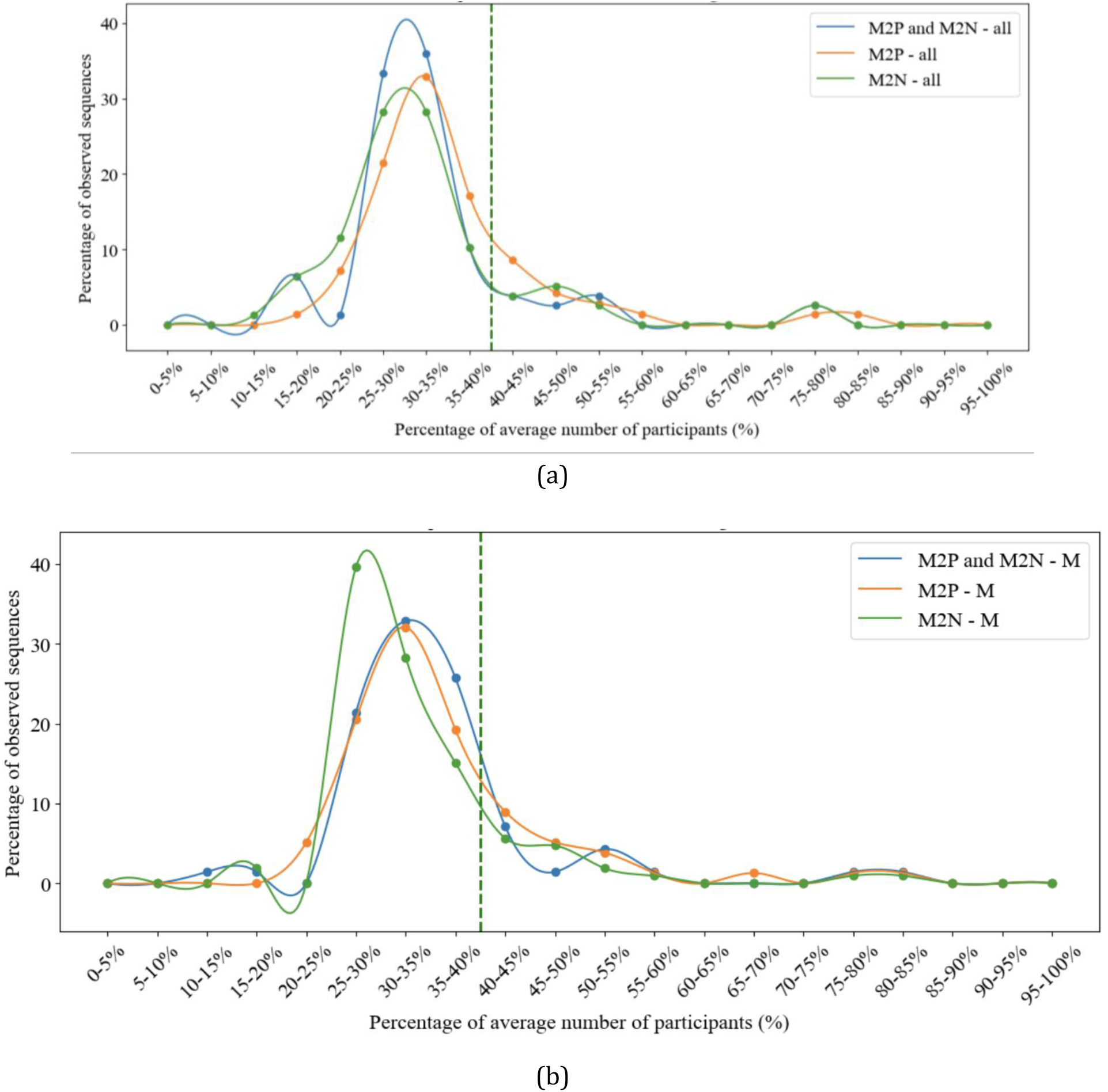
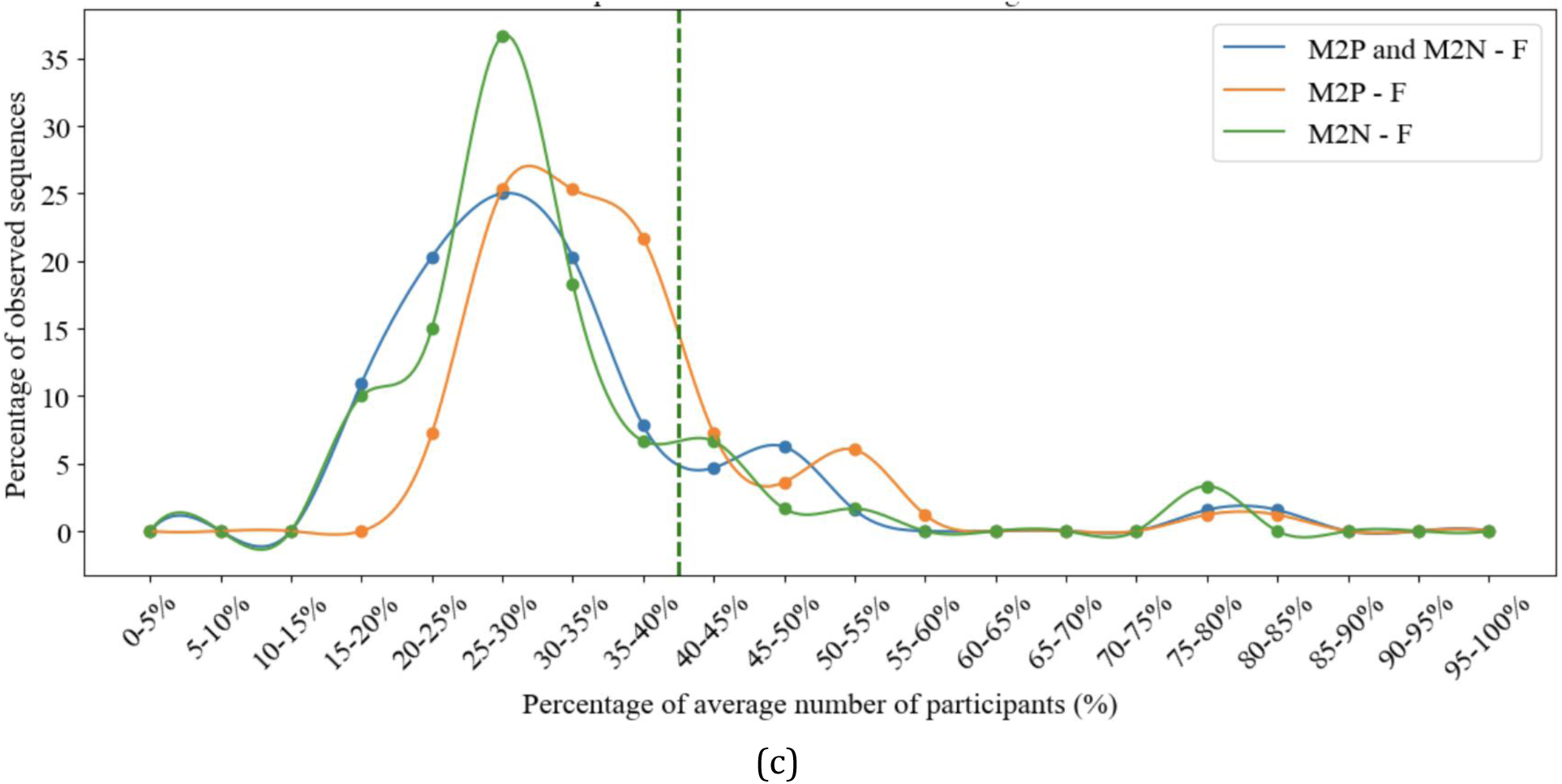
Distribution of the percentage of observed sequences for DCC by average number of participants, considered as percentage for (a) all participants, (b) male and (c) females. Notably, the percentage of females following DCC sequences are more distributed and show considerable variation for M2N and M2P progression from 10-15% to 55-60%, unlike males where most sequences are followed by 20 to 45% of the population.

